# A double-edged sword - Telemedicine for maternal care during COVID-19: Findings from a global mixed methods study of healthcare providers

**DOI:** 10.1101/2020.11.25.20238535

**Authors:** Anna Galle, Aline Semaan, Elise Huysmans, Constance Audet, Anteneh Asefa, Therese Delvaux, Bosede B. Afolabi, Alison El Ayadi, Lenka Benova

## Abstract

**Introduction:** The COVID-19 pandemic has led to a rapid and wide implementation of telemedicine for provision of maternal and newborn health care worldwide. Studies conducted before the pandemic, mainly deriving from high-income countries, showed telemedicine was a safe and cost-effective tool for delivering healthcare under certain conditions. The objective of this study was to document the experiences of healthcare professionals globally with the provision of telemedicine for maternal and newborn healthcare during the COVID-19 pandemic.

**Methods:** We analysed responses received to the second round of a global, online survey of maternal and newborn health professionals, disseminated through professional networks and social media in 11 languages. Data were collected between July 5, 2020 and September 10, 2020. The questionnaire included questions regarding background, preparedness for and response to COVID-19 and experiences with providing telemedicine during the pandemic. Descriptive statistics and qualitative thematic analysis were used concurrently to analyse responses, disaggregated by country income level.

**Results:** Responses from 1,060 maternal and newborn health professional were analysed. Among the sample, 58% reported using telemedicine, with the lowest proportion reported by professionals working in low-income countries (24%). Two fifths of telemedicine users reported not receiving guidelines on the provision of care through technology. Key practices along the continuum of maternal and newborn healthcare provided through telemedicine included online group birth preparedness classes, antenatal and postnatal care by video/phone, setting up a COVID-19 helpline at maternity wards, and online psychosocial counselling. Challenges reported technological barriers, lack of technological literacy, financial and language barriers, lack of nonverbal feedback, and distrust from patients. Maternal and newborn health providers considered telemedicine to be an important alternative to in-person consultations to maintain care provision during the COVID-19 pandemic. However, they also emphasized the lower quality of care and risk of increasing the already existing inequalities in access to healthcare.

**Conclusions:** Telemedicine has been applied globally to address the disruptions of care provision during the COVID-19 pandemic. However, some crucial aspects of maternal and newborn healthcare seem difficult to deliver by telemedicine. Pitfalls of health care provision by telemedicine include exacerbated inequalities in access to care, patient-provider communication problems, and a financial burden for certain healthcare workers and women. More research regarding the effectiveness, efficacy, and quality of telemedicine for maternal health care in different contexts is highly needed before considering long-term adaptations in provision of care away from face to face interactions. Clear guidelines for care provision and approaches to minimising socio-economic and technological inequalities in access to care are urgently needed.

**Summary box:** *What is already known?:*

- Telemedicine is the delivery of healthcare services by healthcare professionals from distance through using information and communication technologies for the exchange of valid and correct information.
- Telemedicine for maternal and newborn health can safely be used to deliver certain components of care in highly controlled settings where the technology is available and accessible to patients
- Telemedicine has been applied rapidly and on a wide scale during the COVID-19 pandemic to replace face to face visits along the continuum of maternal and newborn health care.

*What are the new findings?:*

- Maternal and newborn healthcare providers globally considered telemedicine of benefit during the pandemic and applied it on a wide scale for different aspects of maternal and newborn healthcare.
- The rapid adaptation to provision of care via telemedicine was not optimally supported by guidelines, training for health providers, adequate equipment, reimbursement for cost of connectivity and insurance payments for care provided remotely.
- Healthcare providers reported not being able to reach a substantial group of families by telemedicine and encountered different barriers in providing high quality maternity care by telemedicine due to challenges present worldwide, but more prominent in low- and middle-income countries.

*What do the new findings imply?:*

- Pre-existing inequalities in terms of access to high quality care might have increased by the large scale and rapid implementation of telemedicine during the COVID-19 pandemic in different settings.
- Access to telemedicine for women was hampered by various factors such as internet connection problems, lack of the necessary equipment, digital illiteracy and distrust.
- In-depth research is needed to formalise evidence-based guidelines for the implementation of telemedicine along the continuum of maternal and newborn care as lessons learned for building back beyond the COVID-19 pandemic and also for future emergency preparedness.

## INTRODUCTION

The World Health Organisation (WHO) declared COVID-19 a pandemic on the 11^th^ of March 2020, as a consequence of the more than 118,000 cases spread over 110 countries and the sustained risk of further global spread [1]. The overall response strategy in many countries for fighting the pandemic included early diagnosis, patient isolation, monitoring of contacts, isolation of suspected and confirmed cases, and some extent of lockdown[2]. In this context, the vital role of telemedicine, video consultations in particular, rapidly increased in order to reduce the risk of transmission, especially in settings where insufficient personal protective equipment (PPE) was available for the health workforce. The call for implementing telehealth rapidly has never been louder, especially in high-income countries (HICs) where technological resources are widely available [3–5]. As a consequence, an updated framework for telemedicine in the COVID-19 pandemic for aiding national governments in defining their strategy against COVID-19 was proposed at the start of the pandemic, advising telehealth for all routine care and (suspected) COVID-19 positive cases with mild symptoms [4]. This framework and other variants have been used at a large scale by governments and health systems globally in order to define and implement their public health response to the COVID-19 outbreak while maintaining the provision of essential health services and avoiding the so-called “COVID collateral” [4, 6, 7].

According to the WHO, telehealth is the delivery of healthcare services by healthcare professionals from distance through using information and communication technologies (ICT) for the exchange of valid and correct information [8]. Sometimes the term telemedicine is specifically used to refer to service delivery by physicians only, while telehealth includes service provision by nurses, pharmacists, and other health professionals. In a broader definition, the term telehealth also includes the interactions between health care providers (e.g., delivery of training, team meetings) and the interaction of a patient with technology in the absence of a health provider (e.g. an automated phone line, a health application on a mobile phone)[9]. In this paper, we use the terms telemedicine and telehealth synonymously and interchangeably[8]. A wide variety of telehealth interventions exist today, including the use of mobile phone applications (apps), online health education modules, web portals, wearable devices, text messaging (SMS), and live audio-visual communication[9]. The benefits of investing in telemedicine (sometimes also referred to as m-health or e-health[10–12]) have been discussed in the literature mainly from a public health perspective, documenting the many successes in different health service domains and countries[13–15], ranging from economic efficiency to overcoming distance barriers in remote areas[16]. Recently, the use of telehealth has been expanded drastically during the COVID-19 pandemic due to the necessity to ensure physical distancing as a strategy for slowing down the transmission of the virus, and the limited accessibility of healthcare to both patients and providers [3]. A general consensus seems to have emerged among health experts and authorities that the benefits of telemedicine (especially for non-emergency care) outweigh the disadvantages during the COVID-19 pandemic [3, 13].

Previous epidemic outbreaks and health system disruptive events showed the potential of telemedicine in avoiding further spread of an epidemic disease and maintaining some provision of general healthcare[4]. A mobile app named Ebola Contact Tracing could successfully monitor and contact trace confirmed cases during the Ebola virus outbreak in Sierra Leone and healthcare providers were educated and trained by a virtual tutorial[17, 18]. Studies also explored the use of telehealth in the context of natural disasters and conflicts, with promising results. In the aftermath of two hurricanes in the US in 2017, telemedicine was successfully used by the implementation of free two-way video consultations for victims, although the lack of infrastructure and Wi-Fi access were cited as serious challenges[19]. More recently researchers noted that infrastructure for connectivity is now increasingly available at both ends of the clinical encounter, especially in the UK and United States [4, 5].

Since the start of the COVID-19 pandemic, telehealth has been increasingly implemented in many countries and for the provision of different types of services, due to a lack of alternatives and pressure to maintain health service provision to some extent [20]. Several studies have reported positive results in terms of providers’ and patients’ experiences, and clinical outcomes [21, 22], but the evidence base so far is scarce and covers a narrow range of health services and contexts. For handling confirmed COVID-19 cases with mild symptoms, the maximum use of telemedicine for guaranteeing providers’ and other patients’ safety has not been questioned as a good practice and essential part of reducing the spread of the virus[23, 24]. On the other hand, the necessity and evidence regarding a shift to providing some elements of care through telemedicine for healthy pregnant women and newborns, remains scarce [25].

The evidence on telehealth interventions in maternal and newborn healthcare before the COVID-19 pandemic is mixed. Two recent systematic reviews found that telehealth interventions were associated with improvements in obstetric outcomes, perinatal smoking cessation, continuation of breastfeeding, monitoring of high risk pregnancies and early access to medical abortion services [26, 27]. The included studies also identified a wide variety of telehealth interventions in maternal healthcare, including the use of mobile phone apps, wearable devices, SMS, and live audio-visual communication. However, only studies conducted in HICs and China, a rapidly growing middle-income country, were included in both reviews. Robust evidence from low- and middle-income countries (LMICs) is lacking, mainly due to poor methodical quality of studies and their narrow scope (often focusing on a single application in a specific setting) [28]. SMS support during pregnancy (associated with increased utilisation of healthcare, early initiation of breastfeeding, uptake of recommended prenatal and postnatal care consultations, skilled birth attendance, and vaccination) seemed the most promising and commonly evaluated telehealth intervention in low-income country (LIC) contexts[10, 15, 28, 29]. Also in conflict settings and among migrant women telehealth applications for maternal health have been implemented before the COVID-19 pandemic, where they seem especially useful for providing maternal health education [30–32].

The importance of telehealth for maintaining the provision and use of essential maternal and newborn health services during the COVID-19 pandemic has been highlighted by WHO [25]. In its operational guidance for the COVID-19 context, WHO identified health interventions during the antenatal, intrapartum and postnatal periods as essential services and suggested the use of telemedicine when the technology is available [25]. Evidence regarding its implementation, effectiveness, feasibility, cost effectiveness, and health outcomes are beginning to emerge but remain scarce. Available reports, mostly commentaries and grey literature, have highlighted some challenges and concerns for implementing telemedicine in maternal and newborn health alongside its advantages. The availability of technology and connectivity seem to pose a serious bottleneck, together with high start-up costs and lack of health insurance reimbursement for care provided remotely [33]. Concerns regarding a “digital divide”, meaning increasing maternal and newborn health disparities and inequities as a consequence of access to technology and connectivity, have also been raised [34, 35].

Despite the large efforts to supplement the reduction in the provision of in-person maternal and newborn care by telemedicine during COVID-19, little is known about the actual implementation of these efforts and barriers to its effectiveness as perceived by healthcare providers. This paper documents the findings of a rapid online global survey of maternal and newborn health professionals during the COVID-19 pandemic, focusing on their experience of providing care to pregnant and postpartum women and their newborns using telemedicine.

## METHODS

### Study design

In this is study we present the findings from the second round of a repeated cross-sectional online survey of maternal and newborn healthcare providers. We focus on the application of telemedicine for maintaining the provision of maternal and newborn health care during the COVID-19 outbreak. The survey targeted midwives, nurses, obstetricians/gynaecologists, neonatologists, and other health professionals. An invitation to complete the survey was distributed to those who responded to the first round of the survey, and to other healthcare providers through personal networks of the multi-country research team members, maternal/newborn platforms, and social media (e.g., Facebook, Twitter, WhatsApp groups). Additional details about the study design, sampling and findings of the first round of the survey were published previously [36].

### Questionnaire

A team of international experts adapted the questionnaire used in the first round of the survey in light of the evolving situation of the pandemic. The core structure of the first survey round was maintained and we collected data on respondents’ background, preparedness for COVID-19, response to COVID-19, and own work experience during the pandemic. We additionally aimed to expand our understanding and explore in-depth some of the themes that emerged in the responses received during the first round. Particularly relevant to this analysis, we added a section on the use of telemedicine, whereby we asked participants whether they used technology to counsel or provide care to women or their babies remotely, and if so, what type of services were being provided remotely and whether they had received any guidelines on telemedicine provision. We also asked whether they used telemedicine more than before the pandemic, only started at the beginning of the pandemic or in the same way as before the pandemic.

In open text responses, we asked respondents to share the top three successes and challenges that they experienced using telemedicine. Furthermore, respondents could share their general concerns during the pandemic in an open text box at the end of the questionnaire. The questionnaire was available in 11 languages (English, French, Arabic, Italian, Portuguese, Spanish, Japanese, German, Dutch, Russian, and Kiswahili). The questionnaire is available publicly [37] and the questions relevant to telemedicine are provided in Supplementary file 1.

### Data processing and analysis

We included responses collected between July 5, 2020 and September 10, 2020. We cleaned the 1,331 responses received by removing duplicate submissions (n=14), refusals to participate (n=131), submissions with more than 85% of questions with missing answers (n=46), and submissions from respondents who skipped all the telemedicine questions (n=80). Quantitative and qualitative analysis was done simultaneously in a concurrent design. Quantitative analysis involved producing descriptive statistics (frequencies and percentages) using Stata/SE version 14. The number of health providers using any form of telemedicine was the sum of respondents using telemedicine more than before the pandemic, starting with telemedicine at beginning of the pandemic and in the same way as before the pandemic. All open-ended text responses were translated to English by AG (fluent in Spanish, Portuguese, English, Dutch and French) and by AS (Arabic), with additional assistance from the research team with translating and interpreting responses received in other languages. Responses to open-ended questions were analysed using thematic analysis. Braun & Clarke’s six-phase framework was used during thematic analysis and inductive coding was applied [38]. This framework involves a reflexive process of moving forwards (and sometimes backwards) through data familiarisation, coding, theme development, revision, naming, and writing up. The open-ended responses were read and re-read in order to generate initial ideas. Data were then systematically coded, and the codes grouped to develop broader themes. The last two phases involved refining the themes extracted from the data, adding quotes and double checking if the themes really reflected the respondents’ experiences and perceptions with feedback from the co-author group. Throughout this process, we paid special attention to the context in which the participants’ experiences and thoughts were rooted (i.e., country, position in the team, cadre). We tried to identify common patterns among countries and not to focus excessively on outliers, such as rare practices or challenges. Finally, the continuum of maternal and newborn care was used as a framework for visualising the results [39].

### Ethics

This study was approved by the Institutional Review Board at the Institute of Tropical Medicine in Antwerp Belgium under the number 1372/20. Respondents provided informed consent online by checking a box affirming that they voluntarily agreed to participate in the survey.

### Patient and Public Involvement

No patient or public involvement took place in the design or conduct of this study.

## RESULTS

Table 1 displays the background characteristics of the 1,060 survey participants included in the analysis. Respondents worked in 71 different countries, most commonly in Kazakhstan (n=507), the Democratic Republic of the Congo (n=43), Italy (n=43), Nigeria (n=37), and Japan (n=34). The most common cadres of health professionals in the sample were nurses (29%), midwives and nurse-midwives (25%), and obstetricians/gynaecologists (21%). The majority of respondents identified as females (78%). Respondents were most commonly involved in providing outpatient antenatal care (ANC) (38%), outpatient postnatal care (PNC) (30%), inpatient childbirth care (34%), and inpatient PNC (31%). Three quarters of respondents worked in public health facilities, almost half worked in urban settings (large and small cities), and 22% worked in villages or rural areas.

**Table 1.**
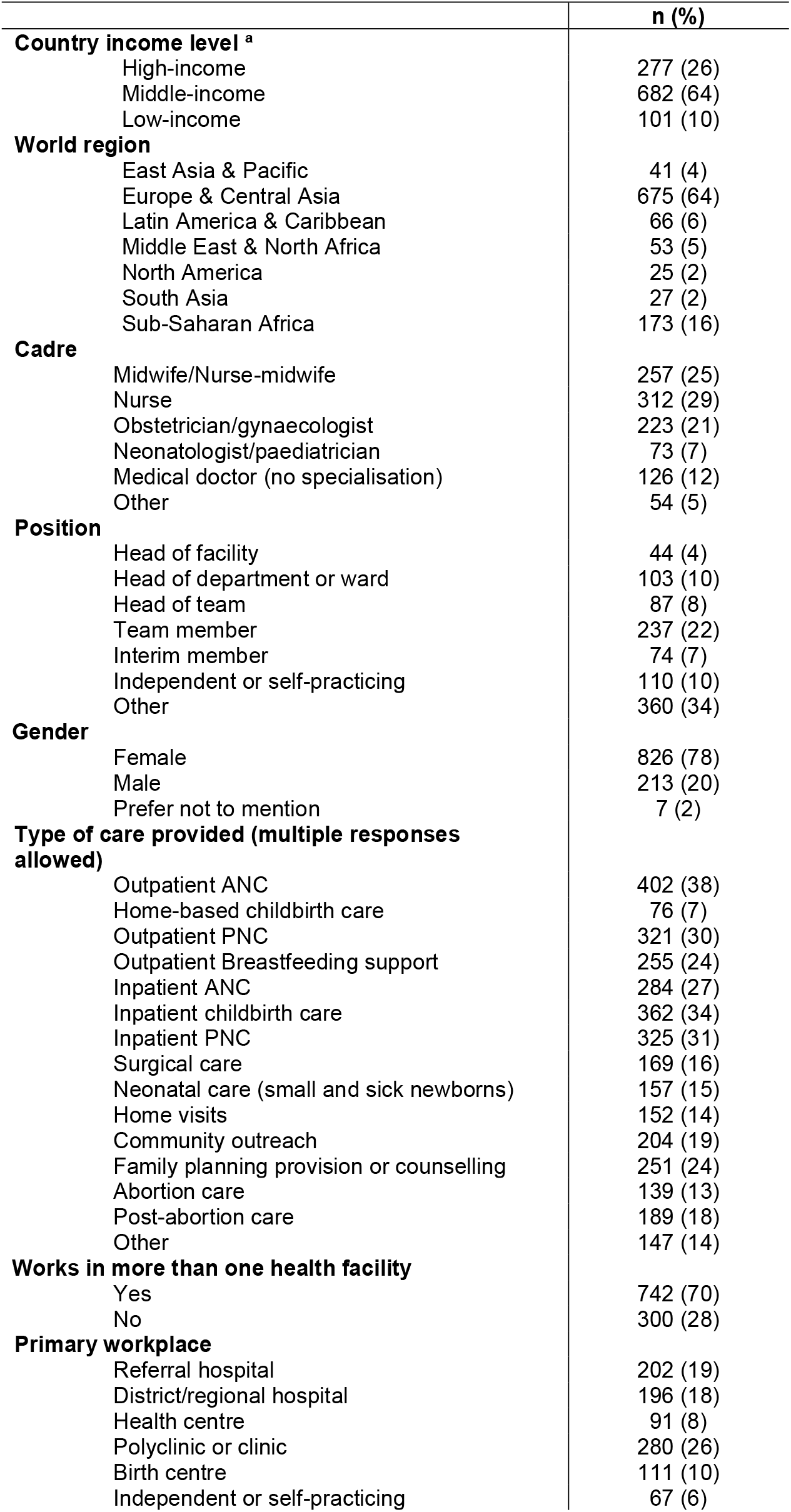
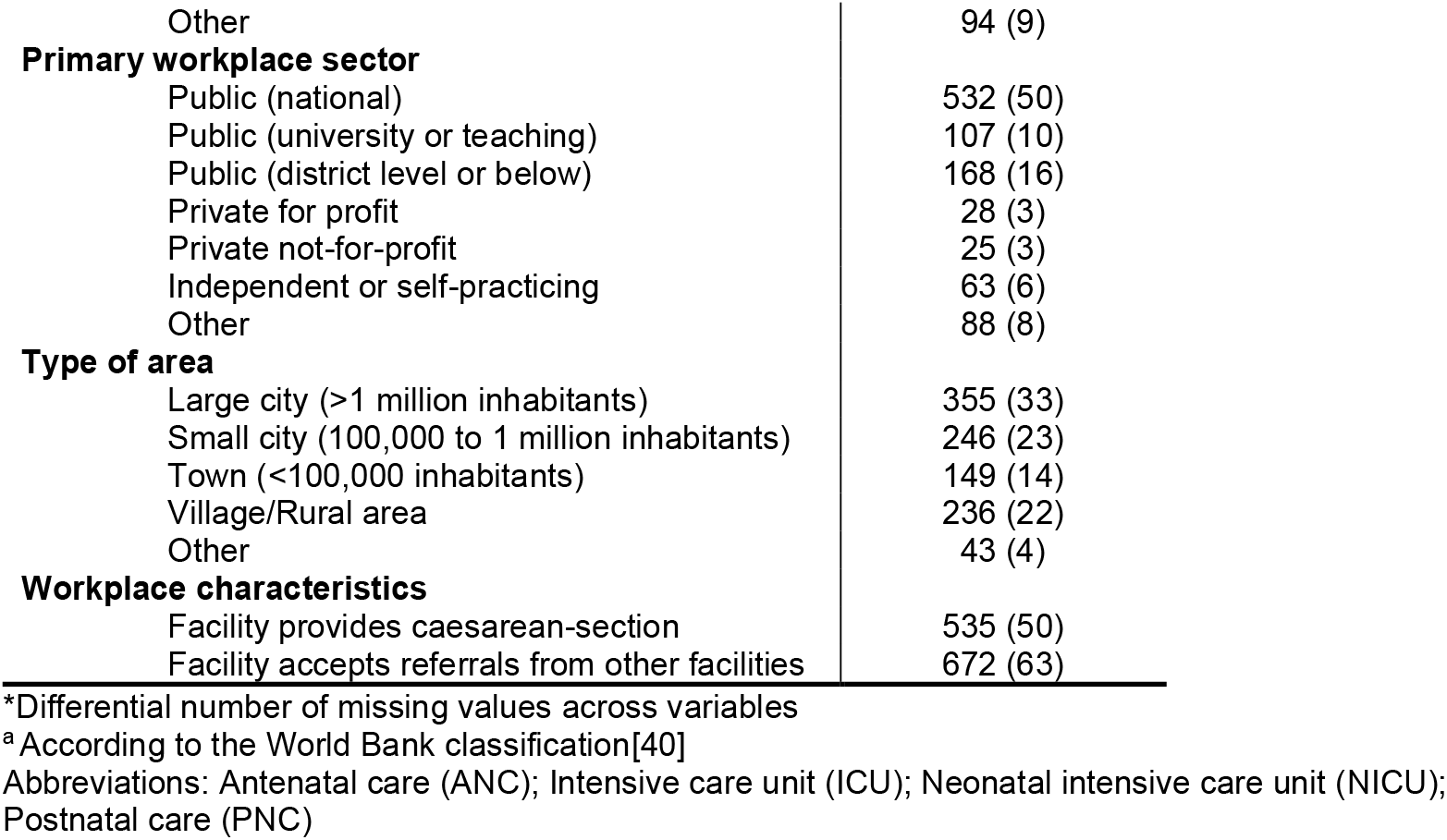
Respondents’ background and workplace characteristics (n=1,060*)

|  | n (%) |
| --- | --- |
| **Country income level <sup>a</sup>** |  |
| High-income | 277 (26) |
| Middle-income | 682 (64) |
| Low-income | 101 (10) |
| **World region** |  |
| East Asia & Pacific | 41 (4) |
| Europe & Central Asia | 675 (64) |
| Latin America & Caribbean | 66 (6) |
| Middle East & North Africa | 53 (5) |
| North America | 25 (2) |
| South Asia | 27 (2) |
| Sub-Saharan Africa | 173 (16) |
| **Cadre** |  |
| Midwife/Nurse-midwife | 257 (25) |
| Nurse | 312 (29) |
| Obstetrician/gynaecologist | 223 (21) |
| Neonatologist/paediatrician | 73 (7) |
| Medical doctor (no specialisation) | 126 (12) |
| Other | 54 (5) |
| **Position** |  |
| Head of facility | 44 (4) |
| Head of department or ward | 103 (10) |
| Head of team | 87 (8) |
| Team member | 237 (22) |
| Interim member | 74 (7) |
| Independent or self-practicing | 110 (10) |
| Other | 360 (34) |
| **Gender** |  |
| Female | 826 (78) |
| Male | 213 (20) |
| Prefer not to mention | 7 (2) |
| **Type of care provided (multiple responses allowed)** |  |
| Outpatient ANC | 402 (38) |
| Home-based childbirth care | 76 (7) |
| Outpatient PNC | 321 (30) |
| Outpatient Breastfeeding support | 255 (24) |
| Inpatient ANC | 284 (27) |
| Inpatient childbirth care | 362 (34) |
| Inpatient PNC | 325 (31) |
| Surgical care | 169 (16) |
| Neonatal care (small and sick newborns) | 157 (15) |
| Home visits | 152 (14) |
| Community outreach | 204 (19) |
| Family planning provision or counselling | 251 (24) |
| Abortion care | 139 (13) |
| Post-abortion care | 189 (18) |
| Other | 147 (14) |
| **Works in more than one health facility** |  |
| Yes | 742 (70) |
| No | 300 (28) |
| **Primary workplace** |  |
| Referral hospital | 202 (19) |
| District/regional hospital | 196 (18) |
| Health centre | 91 (8) |
| Polyclinic or clinic | 280 (26) |
| Birth centre | 111 (10) |
| Independent or self-practicing | 67 (6) |
| Other | 94 (9) |
| **Primary workplace sector** |  |
| Public (national) | 532 (50) |
| Public (university or teaching) | 107 (10) |
| Public (district level or below) | 168 (16) |
| Private for profit | 28 (3) |
| Private not-for-profit | 25 (3) |
| Independent or self-practicing | 63 (6) |
| Other | 88 (8) |
| **Type of area** |  |
| Large city (>1 million inhabitants) | 355 (33) |
| Small city (100,000 to 1 million inhabitants) | 246 (23) |
| Town (<100,000 inhabitants) | 149 (14) |
| Village/Rural area | 236 (22) |
| Other | 43 (4) |
| **Workplace characteristics** |  |
| Facility provides caesarean-section | 535 (50) |
| Facility accepts referrals from other facilities | 672 (63) |

\*Differential number of missing values across variables
<sup>a</sup> According to the World Bank classification[40]
Abbreviations: Antenatal care (ANC); Intensive care unit (ICU); Neonatal intensive care unit (NICU); Postnatal care (PNC)

Among the entire sample, 58% of health professionals reported using some form of telemedicine (Figure 1). This includes all those who used telemedicine more than before the pandemic, in the same way as before the pandemic, and those who started using telemedicine since the beginning of the pandemic. Three quarters of respondents from LICs were not using telemedicine at all at the time of their response, compared to 41% of those working in high-income and 24% in middle-income countries. The percentage of health care providers introducing telemedicine since the beginning of the pandemic was higher among those who work in high- and middle-income countries (18% and 16%, respectively) compared to low-income countries (1%).

**Figure 1.**
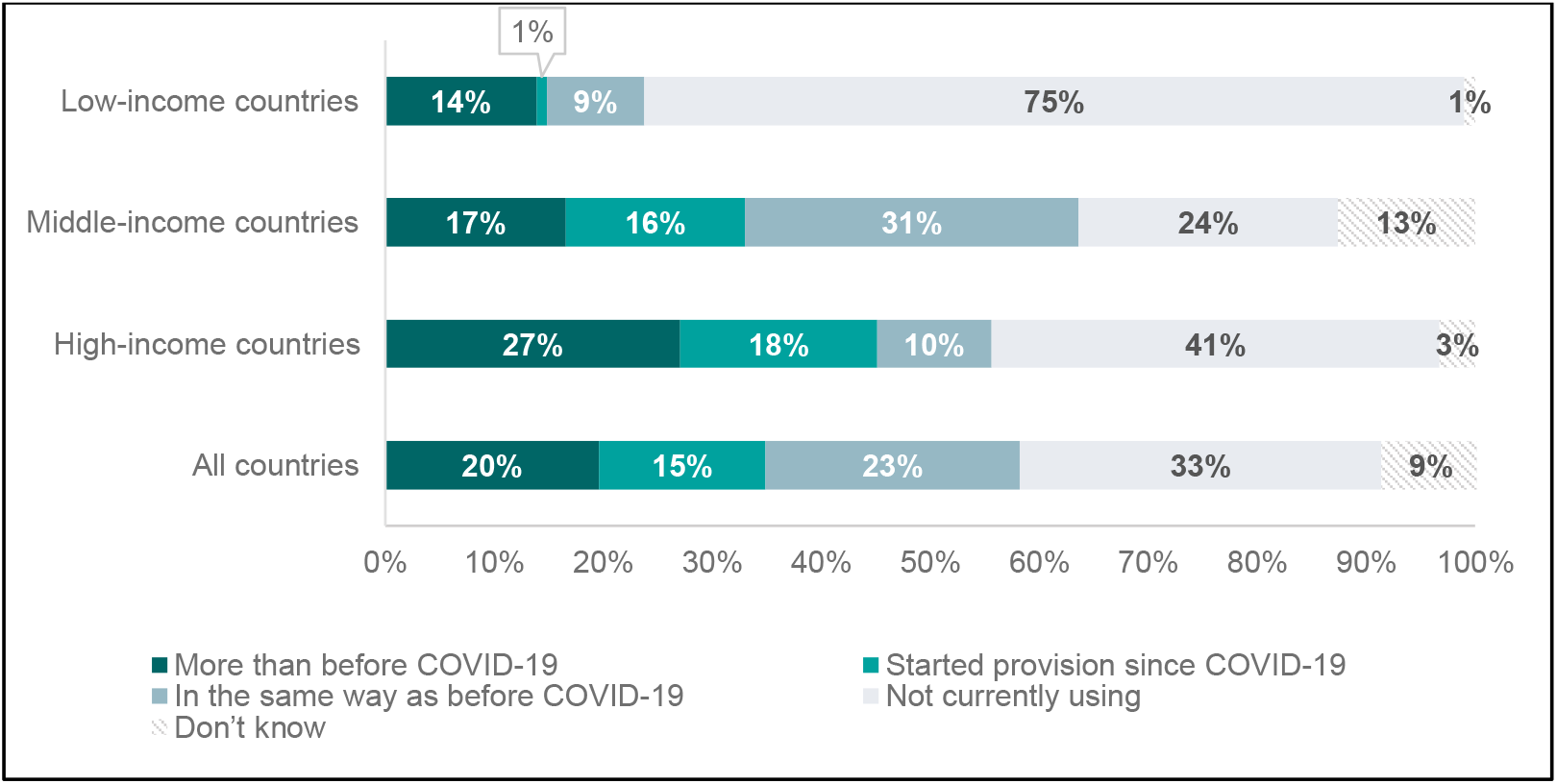
Proportion of respondents currently using technology to counsel or provide care to women or their babies remotely as compared to before the COVID-19 pandemic, by country income level (%, n=1,060)

Among the 612 respondents who provided healthcare using telemedicine, 65% used telemedicine to provide routine ANC, 59% used it to provide childbirth preparation sessions, half provided routine PNC and breastfeeding counselling, 40% provided family planning counselling, and 17% abortion care. Two fifths of respondents using telemedicine reported that they did not receive guidelines on the provision of care through technology.

The main themes identified in the qualitative thematic analysis were the practices and challenges for implementing telemedicine. We identified elements of care along the continuum of maternal and newborn healthcare which were commonly provided using telemedicine (Figure 2). Those were further classified into different telemedicine practices within five broad categories: 1) education and counselling by telemedicine, 2) reducing or eliminating unnecessary personal visits, 3) replacing in-person consultations by telemedicine, 4) setting up hotlines or information lines, and 5) providers connecting to one another by telehealth (Table 2). Furthermore, we found six general themes (not related to a specific component of care) concerning the challenges encountered when providing telemedicine: technological barriers, technological illiteracy, financial barriers, lack of nonverbal feedback, language barriers, and distrust from patients (Figure 2).

**Figure 2.**
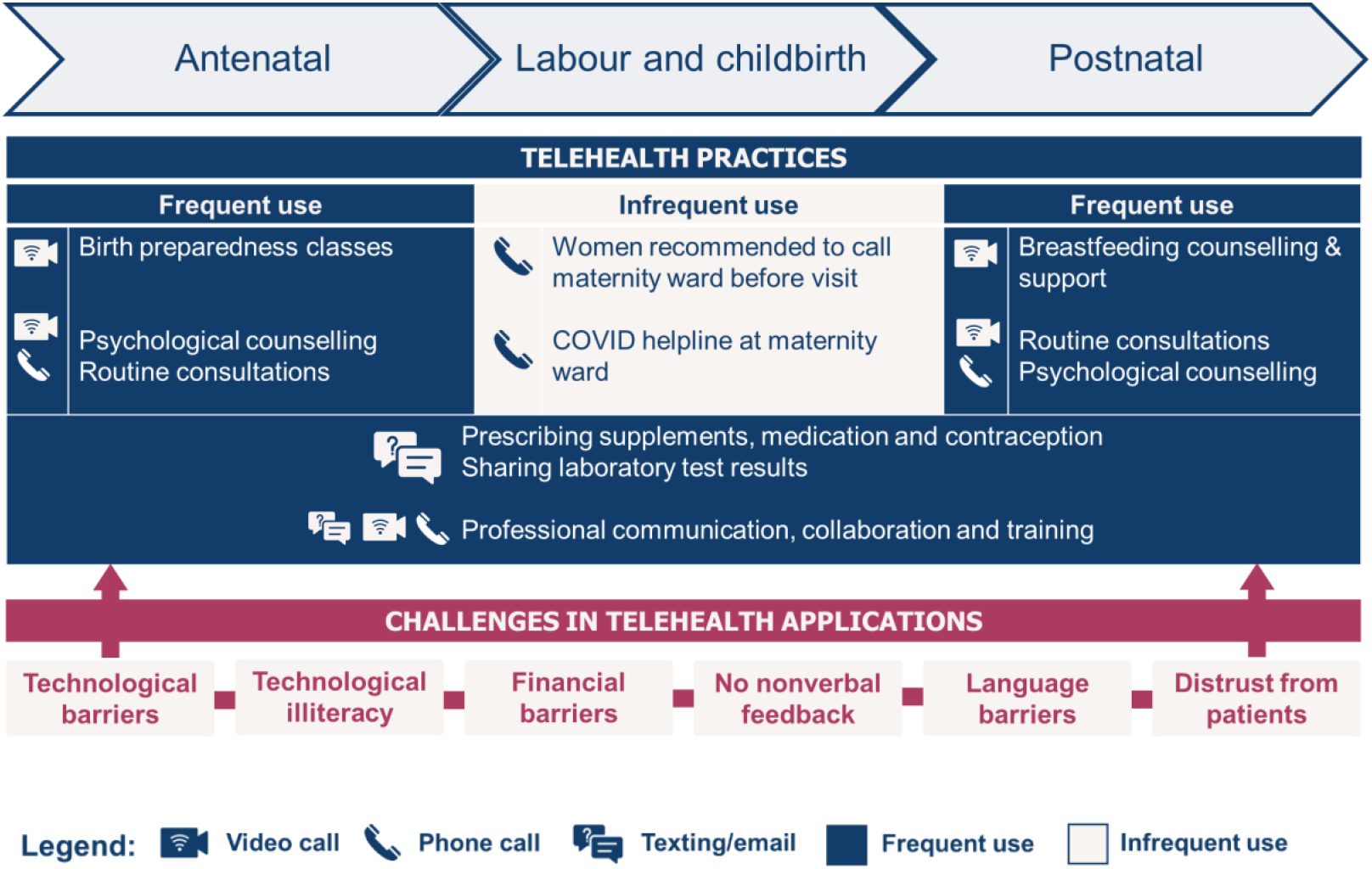
Key types of practices and challenges of providing care through telemedicine along the continuum of maternal and newborn healthcare (n=612) users of telemedicine during COVID-19 pandemic)

**Table 2.** Commonly reported uses of telemedicine in maternal healthcare during the COVID-19 pandemic

| <u>Category</u> | <u>Use of telemedicine</u> | <u>Provider perspectives</u> |
| --- | --- | --- |
| **Education and counselling of women and their families** | Providing birth preparedness classes by video | - Allows ongoing provision of important educational and supportive care.<br>- Convenient because women and their partners can participate and continue with other tasks (caring for other children for example) and do not need transport. |
|  | Postnatal (breastfeeding) counselling and support by video | - During the COVID-19 pandemic women often had less access to support from family and friends, making postnatal support by healthcare providers (in particular midwives) even more crucial. |
|  | Psychosocial counselling by phone/video | - Useful to address the generally increased levels of anxiety during the COVID-19 pandemic.<br>- Effective to provide information and highly demanded by women and their families (especially regarding COVID-19 risks and potential negative consequences for mother and baby if infected). |
| **Reducing or eliminating unnecessary personal visits** | Prescriptions (contraception, medication, medical abortion pill) by WhatsApp/email | - Unnecessary visits to the facility can be avoided by giving prescriptions and/or medications for longer periods than usual<br>- Face to face consultation time can be reduced or eliminated when people receive digital information/prescriptions/lab results. |
|  | Lab results exchange by WhatsApp/email |  |
| **Replacing in person consultations with telemedicine** | Antenatal and postnatal care consultations by phone/video | - Inability to perform clinical evaluation (lab tests, palpation, blood pressure monitoring) is a serious shortcoming.<br>- Lack of facial expression (for example for knowing if information was well understood) is a limitation.<br>- Risk of unequal access to maternal healthcare due to a “digital divide” in the population. |
| **Setting up hotlines or information lines to provide guidance on care seeking** | Women with (potential) signs of labour are recommended to call the maternity ward before going to the hospital | - Unnecessary hospital visits can be avoided by giving correct information and telephone counselling.<br>- Accompanied by the fear and risk of “missing” essential information when giving advice by telephone. |
|  | Institutional hotline for all questions regarding maternity care during COVID-19. |  |
| **Providers connecting to one another to provide better care coverage/quality** | Increased professional communication, collaboration and training<br>- Exchange through WhatsApp/email/phone<br>- Online seminars/training sessions<br>- Online simulations | - Useful for discussing concerns regarding high risk patients<br>- Potential to avoid unnecessary referrals by asking advice from experts<br>- Highly used for interdisciplinary care for pregnant and/or postpartum women suspected or confirmed with COVID-19 |

### Practices

The application of telemedicine along the five broad categories was slightly different according to the context, but they seemed to commonly exist in every participating region. Healthcare providers using telemedicine were in general positive and enthusiastic about its potential. For example, a midwife from Norway commented that *“Telehealth is a brilliant way to have a look and give advice without being in touch*.*”*

Providing online group birth preparedness classes during pregnancy was one of the most popular practices of telehealth mentioned by respondents. They explained that telemedicine was a good alternative to face to face classes because education and counselling is an element of care which does not involve a physical examination. This care can be delivered easily by Zoom or other video conferencing platform/mobile application, and providers also reported it was acceptable to pregnant women. Respondents who mentioned using online group birth preparedness classes mostly worked in HICs, indicating that this switch to remote technologies in the provision of health education, in particular group sessions, might have been less common and/or accessible in LICs.

The second telehealth practice concerned the reduction or elimination of unnecessary personal interactions. For example, healthcare providers used email and/or WhatsApp to send prescriptions and laboratory results to their patients. This did not seem to be a new practice, but was reportedly more commonly performed during the pandemic (as women and their families were discouraged from visiting health facilities unless absolutely necessary) and easily accepted and implemented by providers and patients. Medical abortion was an example of a service mentioned within this theme. A doctor from Cameroon described how he assisted women with an abortion, by providing a medical prescription and cervical preparing agent remotely, to reduce the time spent with the patient in person: *“One of the biggest successes of telemedicine is preparing women with abortion remotely for surgical aspiration. I come to perform it when ready*.*”* He mentioned this telehealth practice was routine before the pandemic.

Third, providers commonly reported the application of telemedicine to replace in person consultations. Practices that fall under this theme include conducting antenatal and postnatal consultations through video/phone calls (mostly WhatsApp) instead of in person visits in the health facility or at the woman’s home. Contrary to the exchange of prescriptions and lab results, this seemed an almost entirely newly introduced application of telemedicine. Respondents and patients in some countries were not able to access health facilities due to strict curfews, lockdowns, public transport bans and closures of health facilities, making face to face consultation almost completely impossible. Telehealth was described as a solid alternative to compensate the lack of face to face consultations to some extent, although with limitations. A nurse-midwife from Uganda commented that *“Through phone calls to postnatal mothers on breastfeeding, cord care, and thermal care, babies have survived although they were not able to access the health facilities for postnatal care”*. Healthcare providers reported that their inability to perform physical examinations (such as foetal heartbeat monitoring, blood pressure measurement, or assessing fundal height) was one of the most important shortcomings of antenatal and postnatal consultations through video/phone calls. The practice of remote blood pressure screening or foetal heartbeat monitoring by women themselves (self-monitoring) was not reported by respondents, indicating that this was not a common practice in our sample. Importantly, respondents reported cancelling some antenatal and postnatal consultations without replacement by any form of telehealth, especially for pregnancies considered low-risk. Currently, the WHO recommends eight ANC contacts for a low-risk pregnancy [41]. Some providers reverted to a reduced number of ANC visits by explaining that they returned to the pre-2016 WHO recommendation of four ANC appointments [42]. A midwife from Kenya explained that conducting fewer ANC consultations was done in order to reduce the risk of infection during an ANC visit: *“Overall, we have less consultations because we give women less appointments so that we reduce the risk of being contaminated during consultation”*.

Respondents reported that care during labour and delivery was continued in person during the COVID-19 pandemic. Women continued to be advised to give birth in health facilities. Nevertheless, many healthcare providers perceived a decrease in the number of facility-based births recorded during the pandemic. The only applications of telehealth for intrapartum care were that women in labour were requested to make a phone call before travelling to the hospital, and a shortened hospital stay to just a couple of hours after birth up to a maximum of 48 hours for a normal delivery. This implied that the first days of postpartum care were almost entirely delivered by outpatient care, which was frequently done by telehealth. Only for the visit scheduled at six weeks postpartum some respondents mentioned they foresaw an in person visit to coincide with the newborn’s vaccination schedule. Respondents also reported that women were more in need of guidance and support during the postpartum period (for example with breastfeeding), partly due to reduced informal support by friends and family.

The fourth telehealth practice was established as healthcare providers tried to respond to what they described as higher levels of anxiety, psychosocial problems, questions and insecurity among pregnant and postpartum women during the pandemic. For this purpose they reported using online consultations, phone calls, and text messages. Some health facilities also temporary installed telephone hotlines to answer patients’ questions. Telehealth seemed to grant healthcare providers, midwives in particular, a feeling of connectedness with and caring for their patients under the difficult circumstances of COVID-19. A midwife from Bangladesh described how telehealth combatted loneliness and even saved lives: “*By providing telehealth first of all women are not completely alone. Besides that, it also has saved lives because I advised women by the phone to come on time for the delivery and provided remote abortion care”*.

Fifth, beyond the application of telemedicine as a means of contact between patients and providers, we found that many providers reported a positive effect of the pandemic on collaboration between health facilities and among healthcare providers. They reported an increase in the use of telecommunication for exchange of information and expertise both among colleagues and at institutional levels. A midwife from Germany explained how also team work improved: “*One of the successes during COVID in my organization were the more frequent team meetings, partially done online, which enabled uniform action against the spread of the virus”*. Healthcare providers also mentioned improved interdisciplinary collaboration by sharing guidelines (mostly COVID 19 health protocols) and updates by WhatsApp/email/phone calls. The restrictions regarding physical training courses also increased their participation in online training modules and simulations.

### Challenges

Among the healthcare providers providing telemedicine (n=612) almost half (n=282, 46%) reported challenges with this mode of service provision in the open text responses. Six broad categories of challenges were identified: technological barriers, technological illiteracy, financial barriers, lack of non-verbal feedback, language barriers and distrust.

#### Technological barriers

Most health care providers seemed to use their own smartphone for providing telehealth services and one of the biggest challenges reported was poor internet connection and/or regular interruptions in connectivity. This was a global problem reported by providers from both LMICs and HICs. As noted by a midwife in the United States: *“Trying to connect with women from rural areas with poor wi-fi service was a challenge”*. In LICs, electricity cuts were also mentioned by several respondents, affecting their ability to provide telehealth, as described by a midwife in Nepal: *“The electricity cuts are bothering the most. Besides, internet fall outs*.*”*

#### Technological illiteracy

Lack of skills to manage the software and devices for conducting telehealth seemed a major obstacle for both women and health providers. While many people might have access to the necessary equipment (such as a smartphone), they are reluctant to use it for telehealth because they do not know how to handle the technology correctly or install the necessary app. A midwife from France noted that: “*Women do not always understand how to use the technology properly*.*”* Also at the provider’s side a midwife from Argentina mentioned a lack of technological skills: *“Many providers do not use telemedicine because learning the skills to do it are a personal responsibility instead of an institutional responsibility”*.

#### Financial barriers

While many providers were very enthusiastic about the use of telemedicine, they also noted that it is not affordable for many of their patients because they lack the financial means to purchase the necessary technical devices. WhatsApp (installed on a smartphone) was mentioned most often as communication medium and also preferred by the healthcare provider, but it became clear that not everyone had a smartphone at their disposal. This was a recurring theme in both LMICs and HICs. A medical doctor in India wrote that *“The use of the phone, SMS and WhatsApp is a success for telemedicine but only 30% of the people have a smartphone”*.

Respondents themselves faced financial burdens from the use of telemedicine; several mentioned the absence of reimbursement of costs they incurred while providing telemedicine (including the provided telehealth care consultation itself and associated internet/phone/data costs). Respondents from LICs particularly reported that there was no standard way of getting reimbursed for providing telehealth consultations. Many patients did not have insurance or insurance companies do not cover the telemedicine consultations. Providers did not know how to invoice the telemedicine consultations as patients were not coming personally anymore. In addition, midwives often had to pay their own internet/phone costs, which was perceived as a serious additional barrier. These financial issues were affecting healthcare providers’ willingness and ability to provide care, as noted by a midwife in Kenya in regard to the cost of mobile data: *“Sometimes I cannot do a follow up of the patients because of lack of airtime”*.

#### Lack of non-verbal feedback

Many healthcare providers reported they could not know if the health information was well received/understood by women because they could not read their facial and/or body expressions. Midwives reported this problem more frequently than obstetricians. A midwife in Nigeria elaborated: *“I don’t like offering remote care. I feel that if you are not seeing the non-verbal cues and facial expression of your patient, you will not truly know if they are ok”*. The relationship and bond between the midwife and a woman was also affected by telemedicine. This was described by a midwife in the United States as follows: *“Technology is a good tool, but does not replace face-to-face conversations, palpating a mom’s abdomen, and listening to the baby’s heart rate in order to form warm, trusting bonds between a patient and the midwife”*.

#### Language barriers

Language barriers were perceived to be more problematic during telemedicine compared to in person consultations. Healthcare providers found it easier to overcome language barriers during in person visits by using body language, which was not feasible virtually. Also, the use of interpreters was sometimes not possible or more problematic for online consultations compared to face to face visits. A nurse-midwife from the United States described how both language barriers and financial barriers hampered access to telehealth: *“Using medical interpreters over the video is a real challenge. Furthermore, our most disadvantaged patients also have limited access to telephone or video”*. This theme was typical among midwives who felt that the personal interaction was part of “being a midwife”: *“My main concern is that we are not at the bedside as midwives traditionally are”*, noted a midwife in the United States.

#### Distrust

Healthcare providers perceived that some patients had little trust in care provided through telemedicine and were reluctant to accept it. One specific problem described by healthcare providers was that they perceived undocumented migrants refused telehealth consultations because they were afraid to be recorded during telemedicine interactions and feared a possible prosecution. A midwife in the United States explained: *“Many of my patients are not documented or are in the U*.*S. temporarily, and are thus reluctant to participate in video and telephone visits due to well-grounded fears of information being recorded or listened to by government agencies*.*”*

## DISCUSSION

This study used a rapid global online survey to understand the care process adaptations used by more than a thousand maternal and newborn health providers from over 70 countries during the COVID-19 pandemic. We found that telemedicine was frequently used for various services along the continuum of maternal and newborn healthcare[39], and differently across contexts. The choice of providing telemedicine was often a personal decision of each health professional, rather than a health facility policy or a country guideline. This means that some healthcare providers reported their personal financial costs to be serious barriers to provision. Telemedicine was already practiced to a certain extent before the pandemic by two-fifths of the respondents, but more widely implemented during the pandemic by one in five. Some healthcare providers also introduced telemedicine for the first time during the pandemic. Our findings suggest maternal and newborn health providers in LICs face more severe barriers to implement telehealth practices compared to those in middle- and high-income settings.

In this paper we show that maternal and newborn health professionals adapted the provision of care using telemedicine during the COVID-19 pandemic in many different ways, even within similar settings. The lack of evidence-based consolidated national and universal guidelines together with a legal framework on the usage of telehealth might explain these findings [9, 16]. Two fifths of health care providers using telemedicine did not receive any type of guidelines. Currently, guidelines on telemedicine mostly originate from national medical specialty societies, limiting their transferability to other health domains and contexts (such as the COVID-19 pandemic) [9]. Furthermore, health systems and governments did not seem to be prepared for the rapid evolution of the pandemic and maintaining the provision of maternal and newborn healthcare might not have been their first priority. Only after the first pandemic peak in March 2020 did the first reports of a potential disruptive effect of the pandemic on maternal and newborn care provision start to emerge[7, 43–46], allowing more coordinated action from stakeholders and governments.

Our study showed that several factors played a role in the decision of health care providers to implement telehealth during COVID-19 or not. These factors included a risk benefit assessment, personal preference of the provider and patient, the financial consequences and health status of the women (low versus high risk pregnancies). Some healthcare providers declared that they only saw high-risk women and shifted entirely to telemedicine for low-risk women. Other providers and facilities shifted to the previous recommendation of four focused ANC visits for low-risk pregnancies[41, 42], replacing some of the remaining visits with telemedicine consultations or a hotline in case of emergencies and questions. A recent publication from India described a similar approach hereby advising face to face ANC provision for high risk pregnant women and a reduced number of visits for low risk women, although they also claimed that more robust data is needed to evaluate the effectiveness of their approach[47]. Obstetric care is characterised by unpredictability; women may develop complications throughout their pregnancy, even when they were classified as low-risk [48]. As a consequence, a low-risk profile might change to high-risk rapidly without warning. Such changes in risk status may go unnoticed, given that healthcare providers reported that providing care via telehealth risks losing certain essential information, which might affect the quality of care especially regarding physical exams and nonverbal feedback. Previous evidence showed that receiving fewer ANC consultations than the recommended has a negative effect on maternal and newborn health outcomes[49–51]. Unfortunately, evidence-based guidelines about the ideal number of telehealth visits versus in person visits during pregnancy and postpartum is lacking, together with guidelines on the integration of home-based equipment (blood pressure monitor, glucometer, urine analysis test strips) which could provide important information to increase care quality and detection of possible complications. In depth research will be needed to assist healthcare providers with guidance on how to implement telehealth along the continuum of maternal and newborn healthcare, and ensure the provision of high-quality maternal health services.

Our study showed that many providers experienced serious challenges in organising and conducting teleconsultations. The most important reported challenges when providing telehealth included technological barriers, technological illiteracy, financial barriers, lack of nonverbal feedback, language problems and distrust from patients. Technological barriers varied from internet connection problems to a lack of smartphones and/or other devices at one or both sides of the clinical encounter (providers and patients). While the lack of internet and equipment is most often reported in LICs [52], it appeared to be a global problem according to our data. A large proportion of healthcare providers and patients do not have easy and affordable access to telemedicine equipment and mobile/data networks. Furthermore, recent research has revealed a gender gap in mobile internet use in LMICs with women being 20 per cent less likely to use mobile internet than men [53]. Given that maternal health is primarily directed to women and that majority of maternal health care workers are women, this gender gap will negatively affect the use of telehealth for maternal healthcare. If telehealth is intended to fill the gaps of healthcare provision during periods of disruptions to healthcare supply and utilisation, or even an essential part of the general healthcare system, this access issue will need to be addressed, possibly through government subsidies or grants for the most disadvantaged groups [54].

Technological illiteracy was another problem commonly reported in our study by respondents from all types of countries. Offering education programs, investing in user-friendly software and social outreach programs might all be strategies to reduce technological illiteracy and the hidden digital inequality and health disparities[55] that might come with broader telemedicine use. Financial issues in using telehealth seemed most problematic in LICs for both providers and patients, according to our study. A big share of women cannot afford to buy airtime for consultations and do not have access to (smart) phones. Healthcare providers also reported struggling to afford equipment and airtime. In addition, healthcare providers found it difficult to get paid for telehealth consultations due to a lockdown and/or lack of reimbursement system. In contrast, health providers in HICs often had access to phones and internet offered by their institutions. Furthermore, health insurance is increasingly covering the main part of the telehealth consultations in line with regular care, which makes the financial compensation for telehealth less problematic in these parts of the world [56–58]. Noteworthy, rapidly evolving technological solutions are coming up in both low- and high-income counties (for example by allowing money transfer by SMS or WhatsApp [59, 60]), which might overcome certain barriers for providing telehealth in the future.

Distrust, lack of non-verbal feedback and language problems were reported as barriers to telemedicine in our study. A study in Bangladesh reporting very similar barriers with telemedicine explained this by emphasising that a provider’s physical presence can easily express empathy and compassion non-verbally, while this is much more difficult during a telehealth consultation[61]. A similar concern was reported by midwives in our study who revealed they could not build a warm and trusting bond with the women by telehealth consultations.

Contrary to the challenges of telemedicine, the benefits of telehealth applications in maternal healthcare are well documented. Many studies show shifting to telemedicine for certain aspects of care is equally beneficial as in person care when it comes to health outcomes and patient satisfaction [62–65]. However, it is important to note that these studies mainly derive from the United States and are conducted in a highly controlled setting with adequate equipment for remote monitoring of blood pressure and blood glucose levels, for example [62, 63, 65]. One study in Japan also reported successful telemedicine provision during COVID-19 by documenting their remote ANC consultation procedure that included the mailing of a cardiotocograph and a sphygmomanometer to each pregnant woman’s home for remote monitoring[66]. Unfortunately, these telehealth interventions do not correspond with the global practice of telehealth during the pandemic, where it has been applied in very different ways and under suboptimal circumstances. Furthermore, telehealth is an already dynamic and rapidly evolving field, resulting in additional challenges for in depth monitoring and evaluation of new applications[16]. In our study, telemedicine was mostly performed by conducting video consultations without reports of patients self-monitoring of vital signs, foetal heartbeat or glucose levels. Very little is known about the benefits and risks of introducing this simplified method and scope of telehealth for providing maternal and newborn healthcare and more research is needed to discover its impact on women and newborns’ health outcomes.

Our study suggests that vulnerable groups are at risk of being excluded from telemedicine, perhaps even higher than from routine in-person maternal healthcare. Given that vulnerable groups such as single women, adolescents, migrants, and women of low socio-economic status already face challenges in reaching traditional maternal healthcare services [67–69], it is crucial that shifting to telehealth does not exacerbate these inequalities. It is important to note that even with concrete guidelines regarding the implementation of telehealth, a one-size-fits-all model will not be appropriate. Each country must continually assess which groups of society are vulnerable to exclusion and fairly support those at the highest risk [70].

Our study showed that in general providers appreciated the application of telemedicine and that it was often the only way to “connect” with women, families and their newborns. Midwives reported higher levels of loneliness and depression among both pregnant and postpartum women, which is in line with the first studies in the field of maternal mental health during the COVID-19 era and previous epidemic outbreaks [71–73]. On one hand the virtual meetings, counselling and support by midwives can help women, but on the other they are only partially doing “the job” because of the lack of physical contact and bonding. More research will be needed on how the mental health needs in the perinatal period can be addressed by telehealth during a pandemic or other similar disruptive situations [74, 75].

Lastly, the COVID-19 pandemic seemed to boost the communication and interdisciplinary management of care between healthcare providers by mobile technologies. Our study showed that the ability to reach colleagues and specialists for advice was valuable to healthcare providers during the pandemic, besides receiving up to date guidelines by virtual communication. Furthermore, they explained it could avoid unnecessary referrals between hospitals by soliciting advice from experts by phone. A Cochrane review also showed that mobile health communication between providers probably decreases the time to deliver healthcare, as well as the number of face-to-face appointments[76], both essential aspects of the global strategy to reduce the spread of the COVID-19 virus. It is critical that this improved collaboration and communication be continued after the pandemic, as the benefits and lessons learned will be important to tackling long-standing issues such as communication during the referral process.

### Limitations

Limitations of our study are the sample bias and lack of representativeness, due to the convenience sampling approach. We received few responses from professionals working in lower-level facilities, particularly in LICs, which might itself be related to limited access to the internet in these settings. Our sample might over-represent higher qualified cadres of health professionals in settings with limited use of technology among lower cadres of staff, and under-represent overstretched staff, or those with limited access to internet connection. This is particularly relevant for the objective of this analysis, as we might overestimate the use of telehealth because of the sample that we reached.

Another limitation is the over-representation of healthcare providers from Kazakhstan in our study. This was a result of the proactive dissemination of the survey by the Ministry of Health in Kazakhstan. A sensitivity analysis was conducted, showing Kazakhstan skewed the data from MICs regarding the use of telemedicine because they were overrepresented (in Kazakhstan 67% of respondents used telemedicine versus 52 % in other MICs). However, this mixed-methods analysis does not aim to make a generalisable statement about telemedicine use by country income levels. We rather use the quantitative findings to frame the qualitative data, which was analysed in-depth to explore healthcare providers’ personal experiences with telemedicine. We therefore do not apply any statistical corrections for the sampling. The overrepresentation of Kazakhstani responses were taken into consideration when summarising and interpreting the qualitative data by stratifying responses according to country and income level, and the emerging themes from Kazakhstan were comparable with those from other settings.

## Conclusion

Maternal and newborn health providers considered telehealth to be an important alternative to providing certain health services during the first months of the COVID-19 pandemic. It gave them the possibility to connect with patients and interact with other health professionals without being exposed to the risks of an in person contact, or when facing restrictions to movement. Furthermore, telehealth seems to be less time consuming (with sometimes even an equal financial compensation) and can easily be combined with other duties at home or in the hospital. However, more research is needed regarding the consequences of an extensive telehealth consultation schedule in maternal health during a pandemic or other emergency situations in the long term. Some authors already pointed to the risk of loneliness and depression for women giving birth during the pandemic, where the lack of interpersonal contact during the postpartum period and increased stress levels seemed serious triggers. We believe the negative consequences might go beyond that, taking into account the reduced number of in-person ANC visits and digital inequality that goes hand in hand with providing telehealth. Especially illiterate, migrant, poor, and ethnic minority women appear to be left behind in accessing maternal health by telehealth. More research regarding the effectiveness and efficacy of telehealth for maternal healthcare in different contexts is highly needed before implementing such adaptations in the long term and on a large scale, particularly to avoid an increase in the existing wide inequalities in access to maternal healthcare worldwide.

## Data Availability

The anonymised data are available from the researcher upon reasonable request and after signing a data sharing agreement.

## Abbreviations

ANC: Antenatal Care
APP: Application (typically a small, specialized program downloaded onto mobile devices)
COVID-19: Coronavirus Disease
ECT: Ebola Contact Tracing
HIC: High-Income Country
ICT: Information and Communication Technologies
ICU: Intensive Care Unit
LIC: Low-Income Country
LMIC: Low- and Middle-Income Country
NICU: Neonatal Intensive Care Unit
PNC: Postnatal Care
SMS: Short Message Service
WHO: World Health Organization

## Acknowledgments

We would like to thank the study participants who took time to respond to this survey during the second round, despite the ongoing difficult circumstances and high workload. We acknowledge the Institutional Review Committee at the Institute of Tropical Medicine for providing helpful suggestions on this study protocol, and for the expedited review of this study. We thank all study collaborators, translators, and colleagues who distributed the invitation for this survey and provided suggestions on the questionnaire, including the co-authors of this paper.

## Contributorship

LB conceptualised the study and obtained funding. All authors contributed to the design of the study and development of the survey tool. AG analysed the qualitative data and AS analysed the quantitative data. AG, AS and LB wrote the original draft of the manuscript. All authors contributed to the development of the manuscript, and read and approved the final version. The corresponding author attests that all listed authors meet authorship criteria and that no others meeting the criteria have been omitted.

## Funding

This study was funded by the Institute of Tropical Medicine’s COVID-19 Pump Priming fund supported by the Flemish Government, Science & Innovation. LB is funded in part by the Research Foundation-Flanders (FWO) as part of her Senior Postdoctoral Fellowship.

The funder had no role in the study design, data collection, analysis, and interpretation of data or in writing the manuscript. Researchers are independent from funders and all authors, external and internal, had full access to all of the data (including statistical reports and tables) in the study and can take responsibility for the integrity of the data and the accuracy of the data analysis is also required.

## Competing interests

None declared.

## Data sharing

Anonymised data analysed during the current study will be made available from the corresponding author upon reasonable request.

## Ethical approval

This study was approved by the Institutional Review Committee at the Institute of Tropical Medicine (Antwerp, Belgium) on March 20, 2020 (approval reference 1372/20).

## Patient and Public Involvement

No patient or public involvement took place in the design or conduct of this study. We involved maternal and newborn health professionals, experts in health systems, infectious diseases, infection prevention and control, and maternal health epidemiologists, and public health researchers from various global settings in the design of this study and the survey tool.

## Dissemination to participants and related communities

The authors intend to disseminate this research through social media, press releases, and media departments and websites of authors’ institutions.

## Transparency

The guarantor affirms that this manuscript is an honest, accurate, and transparent account of the study being reported; that no important aspects of the study have been omitted; and that any discrepancies from the study as planned (and, if relevant, registered) have been explained.

## License for publication

The Corresponding Author of this article contained within the original manuscript, which includes any diagrams & photographs within, has the right to grant on behalf of all authors and does grant on behalf of all authors, a licence to the BMJ Publishing Group Ltd and its licencees, to permit this Contribution (if accepted) to be published in the BMJ and any other BMJ Group products and to exploit all subsidiary rights, as set out in our licence set out at: http://www.bmj.com/about-bmj/resources-authors/forms-policies-and-checklists/copyright-open-access-and-permission-reuse.

## Notes

### Competing Interest Statement

The authors have declared no competing interest.

### Funding Statement

This study was funded by the Institute of Tropical Medicine COVID-19 Pump Priming fund supported by the Flemish Government, Science & Innovation. LB is funded in part by the Research Foundation - Flanders (FWO) as part of her Senior Postdoctoral Fellowship.

### Author Declarations

This study was approved by the Institutional Review Board at the Institute of Tropical Medicine in Antwerp Belgium under the number 1372/20.

