## Supplementary File 1 for "A double-edged sword - Telemedicine for maternal care during COVID-19: Findings from a global mixed methods study of healthcare providers"

**Preparedness and response to COVID-19: a global survey of maternal and newborn care providers**

**Round 2 questionnaire – Questions on Telemedicine**

**Your work and experience in light of the COVID-19 outbreak**

| **Q#** | **Question** | **Response** | **Notes** |
| --- | --- | --- | --- |
| 1 | Are you as a health provider currently using technology to counsel or provide care to women or their babies remotely?  This includes phone calls, video calls, texting, etc. | - Yes – in the same way as before COVID-19 pandemic - Yes – more than before the COVID-19 pandemic - Yes – started since COVID-19 pandemic - No - Don’t know | If no or don’t know, skip questions 1.1 - 1.4 |
| 1.1 | If yes, which type of services?  Please select all that apply | - Routine antenatal care - Childbirth preparation - Routine postnatal care - Breastfeeding counselling - Family planning counselling - Abortion care - Other (please specify) |  |
| 1.2 | Did you receive any guidelines or training on how to provide quality care remotely through technology since the beginning of the COVID-19 pandemic? | Yes  No |  |
| 1.3 | What have been the top 3 key successes related to your use of telemedicine? | [free text] |  |
| 1.4 | What have been the top 3 challenges related to your use of telemedicine? | [free text] |  |
